# Comparison of Two National Noise Models: Progress Towards an Integrated Noise Model for Environmental Health Research in the United States

**DOI:** 10.1101/2025.09.23.25336495

**Authors:** Ching-Hsuan Shirley Huang, Edmund Seto

**Author notes:** Correspondence: Ching-Hsuan Shirley Huang, PhD, Data Science Researcher.

## Abstract

Two sound level maps currently exist for the contiguous United States. One was developed by the National Park Service (NPS) using machine learning methods and sound pressure level monitoring data, and the other by the Bureau of Transportation Statistics (BTS) using transportation noise models of roadway, aviation, and rail sources. Developed for different purposes, each has distinct strengths and weaknesses. This study aimed to compare the two models, develop a hybrid model integrating both, and evaluate its performance against field measurements. Linear regression with data from 378 NPS field sites was used to relate the NPS L50 metric to Leq. A positive association was observed, and the resulting regression equation was used to convert L50 to Leq. Comparing BTS 2018 and 2020 with the converted NPS model, we found strong correlation and small bias between BTS years (Pearson’s r = 0.90, Spearman’s rho = 0.88, bias = 0.3 dBA), but larger differences between BTS and NPS, with BTS levels on average ∼6 dBA higher. A hybrid model was created by filling censored BTS areas with converted NPS Leq values. Evaluation against 708 NPS measurements and 757 metropolitan measurements showed good performance (bias = 0.4 dBA, MAE = 5.0 dBA for NPS; bias = −0.5 dBA, MAE = 3.8 dBA for metropolitan sites). Using the hybrid model, we estimated that ∼36.4 million people (11.1% of the U.S. population) are exposed above 55 dB Leq. The hybrid model provides a resource to inform noise-related environmental health research, policy, and planning.

## 1. Introduction

Models of environmental sound have many practical applications in environmental impact assessment because unwanted sound, or noise, is a nuisance, and is associated with numerous human health impacts. Noise contributes to population annoyance, leading to community complaints (Federal Interagency Committee on Noise FICON, 1992; Fidell, 2003; Miedema and Oudshoorn, 2001; Schultz, 1978). Noise is also associated with sleep disruption, increasing the time to fall asleep, potentially causing awakenings and changes in sleep stage, keeping individuals from falling back asleep, and impairment of subsequent daytime functioning due to inadequate compensatory resting required by the body (Hume et al., 2012; Muzet, 2007). Short-term noise exposure can affect catecholamine levels – the body’s chemical stress response – thereby increasing blood pressure, heart rate, and cardiac output (Babisch, 2011, 2003). Over time, habitual chronic exposure to transportation-related noise especially, is associated with hypertension, ischemic heart disease, myocardial infarction, and stroke (Münzel et al., 2014). Noise also adversely affects children through similar pathways by repeated stimulation of the endocrine and autonomic nervous systems. There is evidence that children’s exposure to noise affects their reading comprehension, memory, and performance on standardized academic tests (Stansfeld and Clark, 2015). Noise can also affect speech perception and listening comprehension for children, thereby impacting their learning (Klatte et al., 2010).

In addition to auditory and non-auditory impacts on humans, noise also has broader ecological impacts. Many animals rely upon sounds to communicate and to identify predators and prey (Sordello et al., 2020). For instance, anthropogenic noise from traffic, construction, industry and other sources may cause auditory masking, interfering with many bird species’ ability to identify important signals (detect prey and communicate via complex birdsongs) from noise, particularly when the frequency of noise overlaps with the communication frequencies of bird species (Dooling et al., 2019). This can lead to physiological and behavioral responses that impact upon species fitness, including effects on reproduction, species abundance, changes in movement, and survival (Engel et al., 2024). Anthropogenic noise can also be propagated through water, affecting many aquatic species. Noise can cause sensory interference related to the specific frequencies of hearing and signaling of aquatic mammals, fish, reptiles, and invertebrates, thereby altering their navigation, foraging, feeding efficiency, defenses against predation, and social structuring (Kunc et al., 2016). Noise pollution can affect biodiversity in both terrestrial and aquatic ecosystems (Harding and Cousins, 2022; Turner et al., 2018). Despite growing evidence on the impacts of noise on wildlife, most US federal agencies, such as the highway and aviation agencies continue to focus noise control efforts in mitigating human health impacts (U.S. Department of Transportation Federal Highway Administration, 2025)

However, a few US agencies have a broader mandate to consider noise and soundscape quality. One is the National Park Service (NPS), a bureau under the US Department of Interior, which conducts soundscape studies in order to conserve and restore the acoustical environment, which is crucial to stewardship of parks, wildlife conservation, ecological integrity, and preservation of the visitor experience for enjoyment by current and future generations (Lynch et al., 2011). They collect ambient sound level data through soundscape measurement campaigns as part of their mission to preserve the natural resources of parks and to understand and mitigate potential adverse impacts of noise on the natural and cultural resources of parks (Schulz et al., 2014). Their measurements rely on high-quality ANSI Type 1 sound level meters and typically involve 25 or more days of continuous collection at national park sites. Their assessments result in detailed technical reports describing the soundscape characteristics of parks and include multiple summary metrics, such as the seasonal A-weighted median ambient sound level (L50) and the natural ambient metric (Lnat) both for daytime and nighttime hours. In addition to measurements, their assessments of park sites typically involve the development of gridded ambient sound levels. In 2014, an effort was made by the NPS to compile data collected across many of their measurement campaigns to develop a contiguous US map of existing, natural, and anthropogenic sound pressure levels (Mennitt et al., 2014). Using a random forest machine learning approach, their model estimated median L50 sound pressure levels during typical summer daytime hours across the contiguous US.

Under the US Department of Transportation, the Bureau of Transportation Statistics has also made efforts to assess sound pressure levels across the US (U.S. Department of Transportation, Bureau of Transportation Statistics, 2020). Since 2018, they have released a nationwide map of transportation-related (major roadway, aviation, and rail) noise (only aviation and roadway noise are available for 2016). Their approach utilizes transportation noise models that are accepted for use by the respective transportation agencies to estimate A-weighted equivalent noise levels (Leq). The resulting map serves both research and planning purposes, allowing for tracking of noise trends and population noise impacts, and for informing transportation-related decisions.

The NPS and BTS maps differ considerably in terms of applicable use scenarios, with one focused on characterizing soundscapes while the other focused on transportation impacts, respectively. They have also chosen different noise metrics (the L50 vs the Leq), which are not directly comparable. Moreover, they each have specific limitations that potentially affect broader applications. For example, a problem with the BTS noise maps is that transportation noise modeling results are not reported in areas with noise levels below 45 dBA Leq. So, with this map alone, sound levels farther away from transportation noise sources are not well defined. But, there is an opposite problem with the NPS map, with most of the sound level monitoring data informing the NPS model being collected from national parks which tended to be lower noise level compared to urban and transportation-dense areas. Yet, both maps have been used in recent national population health assessments. For example, the NPS map of L50 was used by Casey et al. to assess disparities in noise exposure (Casey et al., 2017). Their study found evidence of racial/ethnic and socioeconomic differences in noise exposure across the US. In another study, the BTS map of Leq was used by Huang and Seto to assess high annoyance to transportation noise (Huang and Seto, 2024). This study also observed disparities with respect to transportation noise annoyance. Use of the maps can be challenging though. Because both noise maps can be used for research, policy, and planning applications, it is important to compare noise estimates provided by both models, particularly for more populated areas where large national studies are more likely to recruit individuals for health effects research. There is also the opportunity to combine the two maps together, if the two sound level metrics are harmonized. Most noise mapping efforts to date have been conducted at the city scale (Qin et al., 2024; Seto et al., 2007; Tang et al., 2022). While these studies provide valuable insights into urban soundscapes, there remains a need for consistent national-scale mapping to support broader environmental impact assessment and health research. Our study addresses this gap by harmonizing and integrating two existing U.S. national models.

The key innovations and goals of this study were to (1) develop a statistical relationship between L50 and Leq using sound level monitoring data to harmonize the NPS and BTS maps, (2) compare the harmonized NPS sound pressure level map with the BTS transportation noise map (and also compare the two years of the BTS noise map), (3) create a hybrid national map that combines the BTS and NPS maps, and (4) evaluate the performance of the hybrid map based on comparisons to two sound level monitoring datasets. This is the first attempt that we are aware of to integrate the BTS and NPS maps from two different federal agencies. We discuss the respective strengths and limitations of the BTS, NPS, and hybrid maps.

## 2. Methods

Our approach was to compare the two most recent years of the BTS National Transportation Noise Map with the NPS national sound pressure level model. Because the BTS map is based on the 24-hour equivalent A-weighted sound level (Leq) and the NPS national model is based on the summer daytime median sound level (L50), we utilized a large database of sound level monitoring metrics provided by the NPS to estimate the relationship between summer daytime L50 and the annual 24-hour Leq. This relationship allowed us to convert the L50 results from the NPS national model to estimates of Leq, thereby enabling a more harmonized comparison of sound levels between the BTS and NPS products. Finally, we merged both products into a single noise level estimate by overlaying them spatially and utilizing the estimated Leq from the NPS in locations where the BTS product lacked data (i.e., for levels below the 45 dBA threshold of the BTS noise model). Below, we describe the BTS and NPS noise products, the NPS sound level monitoring data, and the statistical methods for our comparison and data merging.

### 2.1 Bureau of Transportation Statistics National Transportation Noise Map

The National Transportation Noise Map was developed by the BTS to facilitate the consideration of transportation-related noise in policy and planning across the United States. The map is based on the modeling of transportation noise using noise models accepted by federal agencies, and includes noise from aviation, rail, and major roadway sources (U.S. Department of Transportation, Bureau of Transportation Statistics, 2020). Model estimates are provided every two years from 2016 to 2022; however, 2016 does not include rail noise. BTS plans to release model results for 2024 and onwards. Briefly, roadway noise is modeled as the Leq noise metric using the Federal Highway Administration’s Traffic Noise Model (TNM) (U.S. Department of Transportation Federal Highway Administration, 2019) with data on annual average daily traffic (AADT) obtained from the Highway Performance Monitoring System (HPMS), which includes counts of automobiles, medium and heavy trucks on both rural and urban major roadway types ranging from interstate highways to minor arterials and major collector roads. Aviation noise is also modeled as the Leq noise metric, using the Federal Aviation Administration’s Aviation Environmental Design Tool (AEDT) (U.S. Department of Transportation Federal Aviation Administration, 2019) which uses data on flight operations for airports with an average of at least one jet departure per day, excluding airports with exclusively military operations, and excluding helicopter operations. Rail noise is modeled as the Leq using the Federal Transit Administration’s Transit Noise and Vibration Impact Assessment Manual to account for rail equipment and horns on railway lines, and the Federal Railroad Administration’s High Speed Ground Transportation Noise and Vibration Impact Assessment Manual for higher-speed rail operations (U.S. Department of Transportation Federal Railroad Administration, 2012; U.S. Federal Transit Administration, 2018). These rail models use input data on passenger and freight operations, transit system, and freight and commuter rail horns. For roadway, aviation, and rail models, Leq noise is estimated for a 30 m receptor grid. Noise level results below 45 dBA Leq are not reported.

### 2.2 National Park Service National Sound Pressure Level Model

The methods used by the NPS to create a geospatial sound pressure level model for the contiguous United States are described in Mennitt et al. (Mennitt et al., 2014). Briefly, they used summer daytime L50 sound levels obtained from sound recordings conducted between 2000 and 2011 at 190 sites located in National Parks across the contiguous United States to fit a random forest tree-based machine learning algorithm informed by variables related to location and topographic, climatologic, hydrologic, landcover, temporal, and anthropogenic features. The random forest model for L50 (dBA) was found to explain 65% of the variation in sound levels with a median absolute deviation of approximately 3 dB in sound level predictions. The resulting model predictions, provided as a rasterized dataset at 270 m spatial resolution, were obtained from the NPS and represent the A-weighted L50 during a typical summer daytime hour. We focused our analysis on the dataset for “existing” ambient sound levels (as opposed to the “natural” sound level dataset, which excludes anthropogenic sounds based on NPS’).

### 2.3 National Park Service Sound Measurement Data

We obtained 550 summary files of sound recordings collected by the NPS between the years 2001 and 2025. These files generally provided season-specific sound level metrics, including hourly L50 and Leq, as well as daytime values (defined by NPS as between 7:00 and 18:59). We excluded short measurements that did not include all 24-hour period. The resulting dataset consisted of 500 unique sites with 380 summer, 111 fall, 170 winter, and 111 spring measurements (i.e., 772 season-site combinations with sites having between 1 to 4 seasons and a mean of 1.5 seasons per site). Of these files, 378 sites reported summer daytime L50 measurements.

### 2.4 Metropolitan Area Noise Monitoring Data

We obtained annual noise monitoring data for five years (2018-2022) for sites surrounding six airports (SFO, LAX, ORD, JFK, LGA, and EWR) in San Francisco, CA; Los Angeles, CA; Chicago, IL; New York City, NY; and Newark, NJ (Seto and Huang, 2025a). The two California-based airports report noise measurements as CNEL (dBA), whereas the remaining airports report measurements as Ldn (dBA). These measurements were collected at long-term monitoring sites operated by the airport noise offices; however, their monitoring networks include community sites as far as 10 miles (16 km) from the airport. The dataset consists of 144 unique sites and a total of 757 annual noise measurements. Because these measurements were summarized as nighttime (or evening and nighttime) penalized noise metrics, they cannot be directly compared to Leq values without penalties. However, with penalties applied, Ldn will always be higher than Leq. Moreover, if we assume a uniform distribution of sound levels across 24 hours, we can estimate that the Ldn will be approximately 6.4 dBA higher than the Leq (**Table S1**).

### 2.5 Statistical Approach for Hybrid Noise Map and Population Exposure Estimation

To assess the relationship between Leq and summer daytime L50, the 24-hour A-weighted average sound level (Leq) was computed from the hourly Leq metrics in each NPS summary file using conventional acoustical log averaging. These were paired with the summer daytime L50 median sound level, which was parsed from the NPS metric file for each site. We used linear regression to assess the relationship between Leq and L50, with Leq as the outcome and summer daytime L50 as the independent variable (Model A). Statistical significance was defined *a priori* as p<0.05. Two additional sensitivity analyses were conducted: (Model B) a linear regression model with Leq as the outcome and L50 across all hours of the day as the independent variable for all seasons, and (Model C) a linear regression model similar to model B but including a categorical variable for season.

The resulting Model A was used to estimate Leq from the L50 values in the NPS National Sound Pressure Level Model. Estimated NPS Leq values were extracted for all pixels with Leq values in the BTS National Transportation Noise Map for 2018 and 2020. Both Parson’s r and Spearman’s rho correlation statistics were used to compare the Leq values from the different products. Additionally, average bias statistics were computed to determine the typical differences in Leq (dBA) noise levels between BTS years and between the BTS years and the NPS model Leq estimates. Correlation and bias statistics were computed nationwide using a regularly spaced sample of every 4 pixels (>200 million pixels), as well as by the largest 20 (by population) Core Based Statistical Areas (CBSAs).

Finally, a hybrid national noise map was created by overlaying the 2020 BTS transportation model with the NPS estimated Leq model. In the hybrid model, the NPS Leq value was used when the BTS value was missing (i.e., below the BTS’ 45 dBA modeling threshold). To evaluate the performance of the hybrid model, we computed average bias, mean absolute error (MAE) and root mean square error (RMSE) between the hybrid model and Leq values from the NPS sound measurement dataset and the metropolitan area noise monitoring dataset. We then applied this to estimate the population exposure to noise using data from the 5-year American Community Survey (ACS) 2018–2022 at the census block group level (U.S. Census Bureau, 2023). The centroid for each census block group was identified and used to extract the corresponding sound level from the hybrid noise model raster surface. Extracted values were then linked to block group demographic data to generate age-specific exposure distributions. All statistical analyses were conducted in R v.4.5.0.

## 3. Results and Discussion

An approximately linear relationship between summer daytime L50 and Leq was observed from the NPS sound monitoring data (**Figure 1**). The linear regression analysis (Model A) of this relationship (**Table S2**) showed a positive and statistically significant association between the two sound level metrics, with a positive intercept term indicating higher average levels than the median levels. While the slope of the relationship was positive, it was less than one, and we observed (**Figure 1**) that at higher sound levels (e.g., L50 ∼ 60 dBA), Leq was approximately equal to L50. It is reasonable to expect L50 to be lower than Leq, as short episodic sound events (e.g., wind, horns, animal calls, loud vehicle exhausts) are likely to elevate the average but not the median. However, as the frequency of these short-term events increases, it is also reasonable to expect them to influence both the mean and median sound levels. While the data exhibited some scatter, the coefficient of determination (R^2^) of the model was 0.68, indicating that a moderate level of variation is explained by this relatively simple linear relationship. Interestingly, the sensitivity analysis that included data for all seasons (Model B) also found a significant linear association between L50 and Leq (**Table S3**), with similar intercept and slope coefficients as Model A. The other sensitivity analysis, Model C, which included a categorical variable for season, did not indicate that season as a significant additional factor in explaining Leq levels.

**Figure 1.**
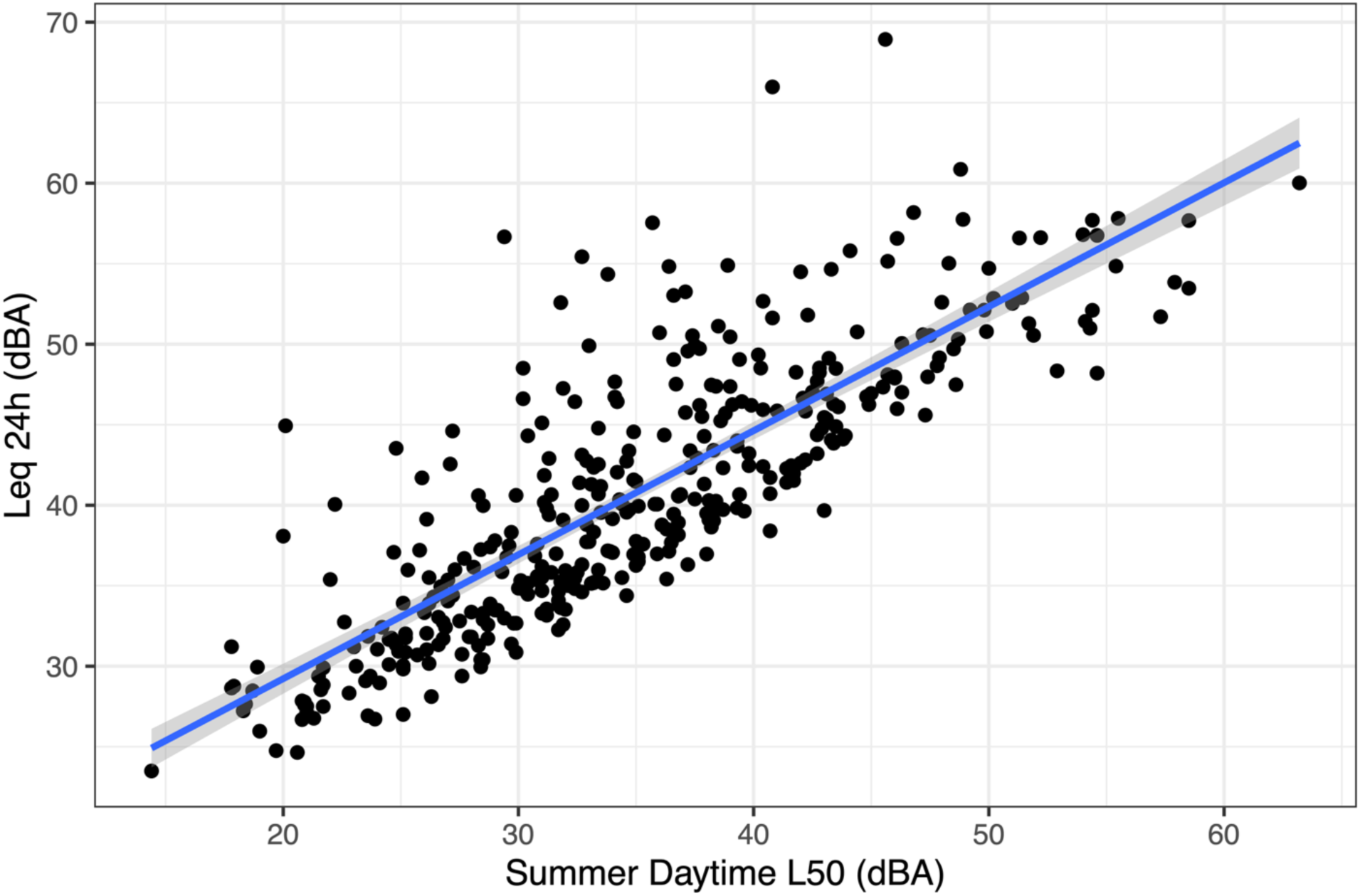
Relationship between summer daytime median L50 sound level and 24-hour equivalent average Leq sound level for NPS sound level monitoring sites. Blue line is the linear model fit with shaded area indicating the confidence interval.

The conversion of the NPS sound pressure level model to Leq (dBA) values is shown in **Figure S1**. Consistent with the applied regression model, we observe that compared to the range of the original L50 values (minimum of 20.1 dBA to maximum of 67.0 dBA), the Leq values were higher at the lower end of the range but approximately the same at the upper end of the range (minimum of 29.3 dBA to maximum of 65.4 dBA). As the L50-to-Leq conversion is only a linear scaling with an offset, the spatial patterns of the original NPS sound pressure level model persist. Notably, we see higher sound levels in urban areas and quieter areas in the Mountain West and Southwest regions of the country.

In contrast, the BTS National Transportation Noise Map shows large areas without modeled Leq noise levels because they fall below the 45 dBA threshold established by BTS (BTS 2018 shown for California state in **Figure S2**). The areas around the main transportation sources used as inputs to the sound dispersion models (e.g., major roads, airport communities, rail lines) can be clearly seen in the map, whereas most other areas are below 45 dBA due to their distance from transportation sources (gray areas in the figure). While the censoring at 45 dBA could be addressed by replacing missing values with the value of 45 dBA, which provides a conservative exposure estimate intended to avoid underestimating health impacts when applying noise exposure-response functions, there are opportunities to utilize the relatively lower yet continuous range of ambient sound levels in the NPS model’s converted Leq values.

Results of the comparisons between the BTS 2018 and 2020 models and the NPS converted Leq model values, including Pearson and Spearman correlations and average bias, are provided in **Table 1**. The two BTS years exhibited high correlation (Pearson’s r = 0.90 and Spearman’s rho = 0.88). However, the BTS transportation noise model was not found to be correlated with the NPS model. Bias statistics indicated that in areas of overlap, on average there was little difference (<1 dBA Leq) between the BTS years. In contrast, on average, sound levels in the BTS model were approximately 6 dBA Leq higher than in the NPS converted values. This suggests a potential underestimation of transportation-related noise in the NPS model, which is largely derived from a machine learning model developed based on measurements collected in national parks, environments generally less impacted by transportation than metropolitan areas and urban cores. We observed this potential underestimation in some transportation-impacted locations. For example, **Figure S3** shows a map of the NPS sound pressure level model with converted Leq values for the Seattle, King County, WA and Chicago, IL CBSAs with a 1 km buffer encircling the respective international airports. The maximal Leq values on these maps do not extend above 65 dBA, levels that would typically be expected around major airports.

**Table 1.**
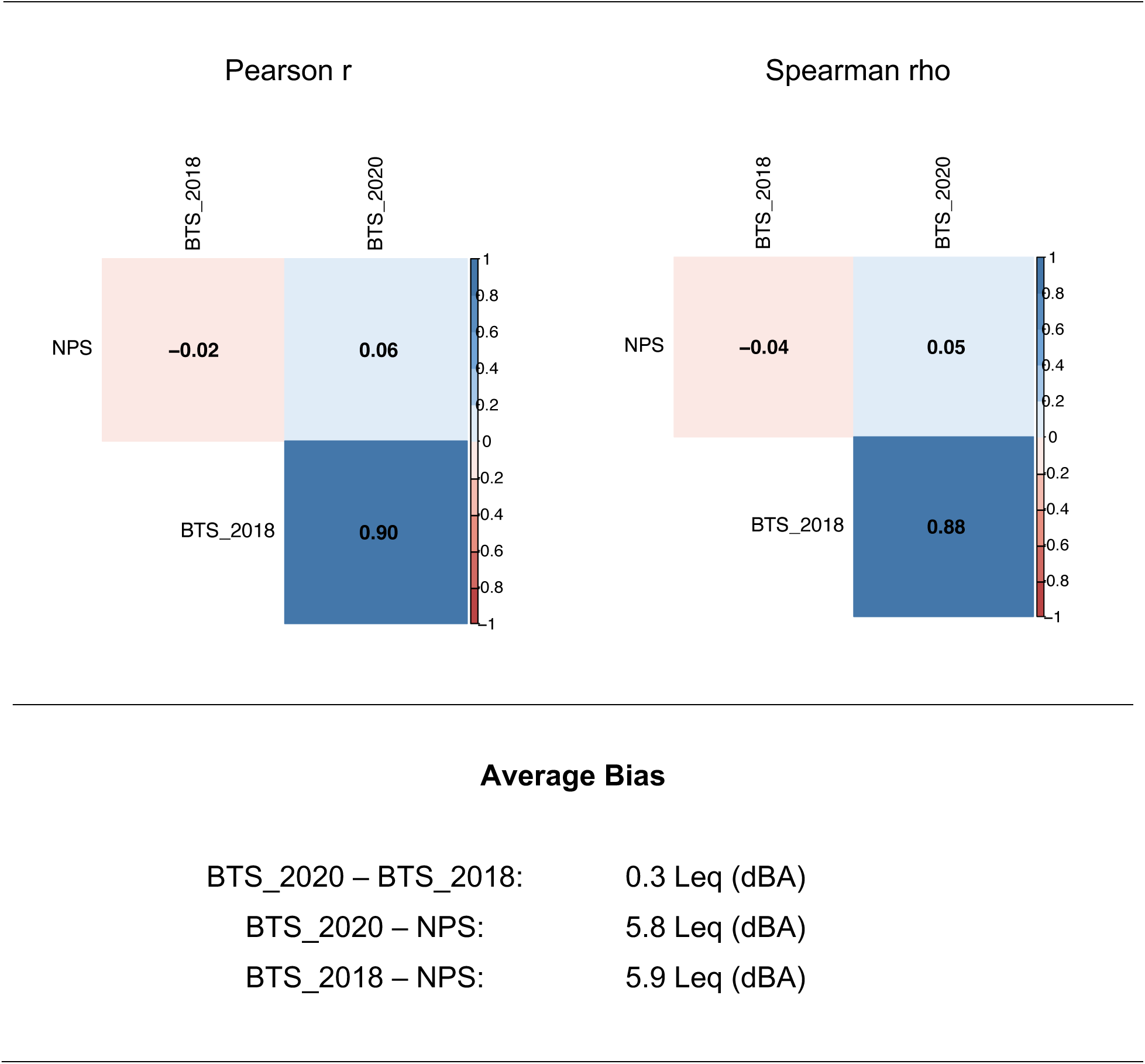
Pearson’s (top left) and Spearman’s (top right) matrices providing correlation coefficients, and average bias statistics (bottom) between BTS National Transportation Noise Model years 2018 and 2020, and the NPS National Sound Pressure Level Model across the contiguous U.S. Color shading of the two correlation matrices indicate the magnitude of positive (blue) vs negative (red) correlation.

Average Bias
|  |  |
| --- | --- |
| BTS_2020 – BTS_2018: | 0.3 Leq (dBA) |
| BTS_2020 – NPS: | 5.8 Leq (dBA) |
| BTS_2018 – NPS: | 5.9 Leq (dBA) |

**Table S4** provides correlation matrices and average bias statistics comparing the two BTS years and the NPS converted Leq model values for each of the 20 largest CBSAs. While there was variation between CBSAs, generally the correlation between BTS years was high in all of the CBSAs (Pearson r = 0.86–0.97), which is consistent with the overall high correlation between BTS years in the nationwide analysis (**Table 1**). Also consistent with the nationwide analysis, we observed relatively weak correlation between the BTS and NPS modeled Leq values (Pearson r = −0.15–0.24 for BTS 2018 vs. NPS). Interestingly, while the nationwide analysis indicated a < 1 dBA mean Leq difference between BTS 2018 and 2020, for some CBSAs (e.g., NY-NJ and Atlanta, GA) the 2018 noise levels were higher than 2020, but for others (e.g., San Diego, CA and Riverside, CA) the 2020 levels were higher than 2018. While transportation levels, such flight operations have been trending higher over time, 2020 was during the COVID-19 pandemic when there were travel restrictions, and thus it is not unusual for noise levels to be lower for some areas during 2020 compared to 2018. Our group’s previous analysis of US Federal Aviation Administration (FAA) flight operations and sound level monitoring data before, during, and after the pandemic found average noise levels to range from 3.0 to 5.4 dBA lower in 2020 compared to peak levels before the pandemic (Seto and Huang, 2025a).

The hybrid model of Leq (dBA) combining the BTS National Transportation Noise Map for year 2020 and the NPS model’s converted Leq values is shown in **Figure 2**. It retains the characteristics of both BTS and NPS models. Notably, urban areas still exhibit elevated sound levels and high sound levels are also observed for areas around transportation-related sources. The range of Leq values includes the high noise levels >60 dBA from the BTS map that coincide with transportation-attributable noise, which are not captured by the NPS L50-to-Leq converted map. However, missing values in the BTS map that are < 45 dBA are now filled in with the NPS Leq values that extend just below 30 dBA. Compared to previous NPS model results for the Seattle and Chicago areas, the hybrid model results do show elevated noise levels > 65 dBA Leq around the airports and other major transportation sources (**Figure S4**).

**Figure 2.**
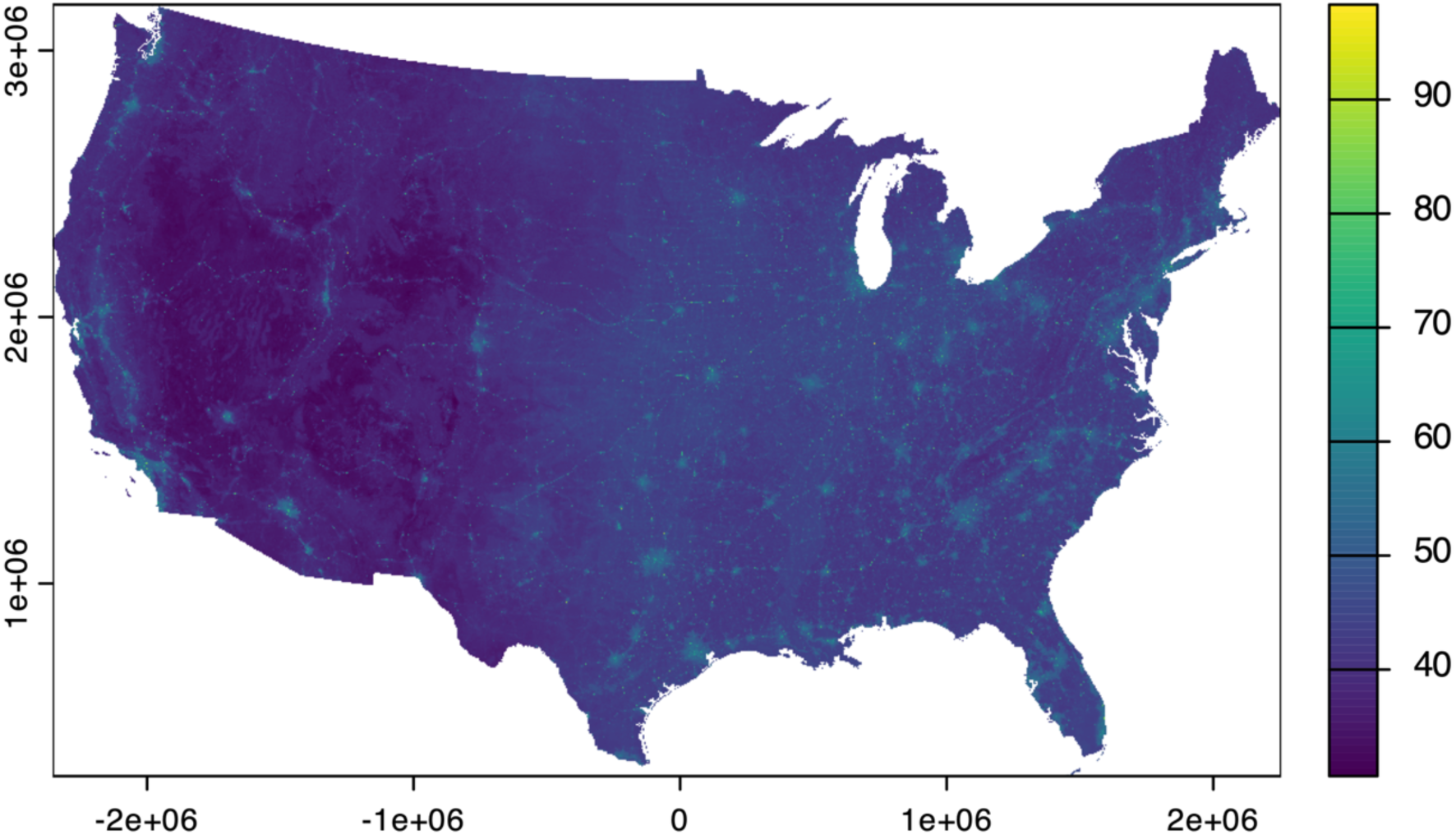
Map of the hybrid model merging the BTS National Transportation Noise Map 2020 with the Leq (dBA) values estimated from the NPS National Sound Pressure Level Model.

To evaluate the performance of the hybrid model, we compared the modeled Leq values to the NPS sound measurement dataset and the metropolitan area measurement dataset (**Table 2**). After excluding measurements with less than 24 hours and sites outside of the contiguous US, the NPS dataset consisted of 708 Leq (dBA) measurements. We found the average bias to be small at 0.4 dBA Leq and the RMSE to be 6.5 dBA Leq. The relatively larger RMSE compared to the average bias suggests that both positive and negative errors exist in the model and approximately cancel each other, resulting in a slight overprediction of <1 dBA compared to NPS field measurements.

**Table 2.** Average bias and RMSE performance statistics for evaluation of the hybrid model Leq (dBA) against the NPS and metropolitan area sound level monitoring datasets.

| Dataset | N | Average Bias<br>(dBA) | MAE (dBA) | RMSE (dBA) |
| --- | --- | --- | --- | --- |
| NPS sites | 708 | 0.4 | 5.0 | 6.5 |
| Metropolitan area<br>sites | 757 | -0.5 | 3.8 | 4.9 |

Because the NPS field measurements tended to be in national parks, we also evaluated the performance of the hybrid model using the separate metropolitan area measurement dataset, which consisted of 144 unique sites and a total of 757 noise measurements. As described in Methods and **Table S1**, because these are penalized noise metrics, they were adjusted downward by 6.4 dBA before comparison to the hybrid model. Average bias and RMSE statistics are provided in **Table 2**. We found an average bias of −0.5 dBA Leq, an MAE of 3.8 dBA, and an RMSE of 4.9 dBA Leq, which indicates a slight overprediction of <1 dBA on average and a lower RMSE compared to performance at NPS field measurement sites in the assessed metropolitan areas.

Because the hybrid model was developed using the BTS 2020 dataset, we conducted an additional sensitivity analysis comparing its performance against metropolitan area field measurements collected in 2018–2019 (N = 265), 2020 (N = 136), and 2021–2022 (N = 243). We observed better performance of the hybrid model for 2018–2019 (average bias = 0.4 dBA; MAE = 3.4 dBA; RMSE = 4.5 dBA) and for 2021–2022 (average bias = −1.7 dBA; MAE = 3.7 dBA; RMSE = 5.0 dBA) than for 2020 (average bias = −3.5 dBA; MAE = 4.3 dBA; RMSE = 5.6 dBA). Studies have observed considerably lower transportation activity during the COVID-19 pandemic, which contributed to reduced transportation-related noise in 2020 (Aletta et al., 2020; Asensio et al., 2020; Čurović et al., 2021; Seto and Huang, 2025a; Walker et al., 2021; Yu et al., 2023; Zambon et al., 2021). The large negative average bias (i.e., field measurements lower than model predictions) for 2020 data is consistent with the 3.0 to 5.4 dBA lower noise levels observed during the pandemic compared to pre-pandemic years (Seto and Huang, 2025a). Because the performance metrics suggest that the hybrid model reflects conditions both before and after the pandemic better than the 2020 pandemic year, the hybrid model may better represent the more generalized noise situation than the 2020-specific situation.

Using these distributions, we further estimated the number of individuals exposed to different noise level (**Table 3**). Across all age groups, the distribution of exposure was similar. Overall, there were approximately 36.4 million people (representing 11.1% of the total population; **Table S5**) exposed to above 55 dB Leq, the threshold defined by the U.S. EPA community noise protection guideline for outdoor activities (U.S. Environmental Protection Agency, 1974). For comparison, the European Union commonly applies a 55 dB Lden benchmark as the reporting threshold for environmental and transport noise exposure (Lambert, 1994; Murphy and King, 2010). Because our model reports Leq rather than Lden, these estimates are likely conservative, as Lden incorporates additional evening and nighttime penalties and therefore yields higher exposure values for the same environment.

**Table 3.** Estimated number of individuals (n) in each age category by sound level exposure (dBA Leq) using the hybrid noise model.

| dBA Leq | All Ages | Age Categories, n |  |  |  |  |  |  |  |
| --- | --- | --- | --- | --- | --- | --- | --- | --- | --- |
|  |  | under 5<br>years | 5 to 17<br>years | 18 to 24<br>years | 25 to 44<br>years | 45 to 64<br>years | 65 to 74<br>years | 75 to 84<br>years | 85+<br>years |
| <35 | 316,166 | 17,780 | 54,224 | 29,347 | 76,915 | 79,289 | 36,420 | 17,572 | 4,619 |
| 35-39 | 4,131,339 | 205,963 | 657,952 | 287,799 | 929,179 | 1,121,292 | 589,636 | 262,481 | 77,037 |
| 40-44 | 41,826,971 | 2,201,521 | 6,847,109 | 3,201,886 | 9,432,289 | 11,728,586 | 5,163,755 | 2,453,410 | 798,415 |
| 45-49 | 94,326,184 | 5,344,572 | 16,252,856 | 8,389,573 | 23,182,027 | 24,650,970 | 9,722,295 | 4,861,729 | 1,922,162 |
| 50-54 | 151,798,740 | 8,987,313 | 24,773,531 | 15,036,707 | 42,255,526 | 37,207,266 | 13,646,577 | 6,860,550 | 3,031,270 |
| 55-59 | 31,281,655 | 1,813,714 | 4,479,839 | 3,573,057 | 10,137,990 | 7,119,080 | 2,436,677 | 1,187,923 | 533,375 |
| 60-64 | 2,947,993 | 172,146 | 457,074 | 338,223 | 887,088 | 681,538 | 242,828 | 117,621 | 51,475 |
| 65-69 | 1,176,070 | 70,397 | 173,806 | 125,348 | 371,467 | 270,611 | 96,350 | 46,987 | 21,104 |
| 70-74 | 508,135 | 27,736 | 78,137 | 50,192 | 151,297 | 125,769 | 42,397 | 23,483 | 9,124 |
| 75-79 | 291,987 | 15,634 | 44,051 | 27,585 | 90,174 | 70,317 | 24,205 | 13,352 | 6,669 |
| 80-84 | 128,482 | 6,631 | 19,355 | 16,240 | 42,393 | 28,066 | 9,943 | 3,733 | 2,121 |
| 85-89 | 20,798 | 1,130 | 2,185 | 3,360 | 7,081 | 4,357 | 1,586 | 838 | 261 |
| 90+ | 15,950 | 1,081 | 2,477 | 1,611 | 4,273 | 4,365 | 1,356 | 657 | 130 |

This study represents one of the first attempts to compare and integrate two national US noise models: the NPS model, developed through machine learning methods, and the BTS model, developed through transportation noise modeling. The resulting hybrid model addresses inherent limitations in the original models, including their use of different noise metrics, the reliance on data collected primarily from natural soundscapes in the NPS model, and the focus solely on transportation-related noise in the BTS model. We observed potential underestimation of transportation-related noise in the NPS model. Conversely, the BTS map exhibited large areas of censored (missing) values due to its 45 dBA modeling threshold. We found relatively small differences in predicted noise levels between the BTS model years 2018 and 2020, compared to the larger differences observed between BTS and NPS models. Using field monitoring data to develop a relationship between L50 and Leq, and conversion of the NPS L50 modeled values to Leq allowed for the use of the NPS model to fill <45 dBA gaps in the BTS map. This resulting hybrid map tended to perform well when evaluated against NPS sound measurements and additional metropolitan area sound measurements. However, we observed that the reduction in noise that occurred during the pandemic year (2020) may not be fully represented in the hybrid model despite the inclusion of noise levels from the BTS year 2020 transportation noise map. But the hybrid model was found to perform well when evaluated against pre- and post-pandemic noise measurements. Thus, the hybrid model is probably a more appropriate representation of longer-term average noise levels rather than a year-specific model.

This hybrid model has numerous potential practical uses. Because both NPS and BTS are continuing to update their models, there are opportunities in the future to integrate the two in future updates of the hybrid model to inform noise-related policy, urban and transportation planning, and environmental, ecology, social, economic, and public health-related studies. Our group is particularly interested in exposure sciences research, such as the assignment of environmental noise exposures to human populations based on their residential address locations. Previous studies have utilized the L50 sound estimates from the NPS model for such research (Casey et al., 2017), and may benefit from use of the hybrid model due to its improved characteristics. In addition to applications in exposure assessment and planning, detailed noise maps may also support interpretation of community complaint data, which are increasingly used in environmental impact studies (Pinsonnault-Skvarenina et al., 2024; Ramphal et al., 2022). Integrating noise modeling with complaint-based approaches could improve understanding of how populations perceive and respond to noise.

One key improvement of the hybrid model is the use of Leq rather than L50, which enhances comparability with noise-related health research conducted internationally. For example, although Lden and Lnight are primarily used in the European Noise Directive, and Ldn is used by the FAA and many state and local noise ordinances, these metrics are ultimately aligned with the concept of Leq average noise levels, rather than the median. In fact, many exposure-response functions used to estimate noise-related health impacts are based on average or penalized average noise levels rather than L50, including the classic Schultz high-annoyance curve and more recent curves developed by the US Federal Interagency Committee on Noise (FICON) (Federal Interagency Committee on Noise FICON, 1992; Fidell, 2003; Schultz, 1978). Variations nevertheless exist in the use of certain metrics in the US. For example, CFR Part 150 for compatible land use around airports relies on assessment of the Ldn. Other agencies such as the US Housing and Urban Development also relies on Ldn in its consideration of “a decent home and suitable living environment for every American family” (Office of Community Planning and Development, 2014). Some states, such as California have adopted their own metric, for example, CNEL (a day-evening-nighttime penalized noise metric similar to Lden) for noise policy, including at airports. Individual cities (e.g., Seattle, WA) may primarily rely on Leq for their noise control ordinances, although these often emphasize reduced sound levels during nighttime hours (City of Seattle, 2025).

While the focus of this study was on comparing and integrating the two models, our analysis benefited from the use of a large number of sound level measurements to develop the L50-to-Leq relationship and to evaluate the performance of the hybrid model. As there is no single agency that is responsible for compiling noise measurements for the US, these data were not readily available and had to be painstakingly requested, processed through quality checks, and integrated with site location data. In some cases, data used for this project had to be manually extracted from technical reports. There were over 1,400 measurements used in this study. As we continue to compile more noise monitoring data, there are opportunities to move beyond integration of two existing models to develop new machine learning models that more accurately reflect spatial and temporal variations in noise across the US.

Currently, this work is limited in its consideration of standard noise indicators, such as the L50 and Leq for national noise modeling. However, recent research in the US, such as the FAA’s Neighborhood Environmental Survey (NES), suggests that people may perceive noise differently than before (Miller et al., 2021; Seto and Huang, 2025b). Findings from NES indicate that for a given Ldn sound level, the percentage of people reporting high annoyance may be much greater than previously estimated by the Schultz or the FICON curves. Temporal, frequency-related, and non-acoustic factors may contribute to the changing attitudes towards noise and the health impacts of noise exposures (Guski, 1999; Guski et al., 2017). These different characteristics are difficult to capture with a single noise indicator. Future nationwide noise modeling efforts may benefit from the consideration of other indicators, such as metrics that capture the distributional characteristics of sound (e.g., L10, L50, L90 percentile measures), spectral frequency characteristics (e.g., lower frequency sounds, fundamental vocal frequency range where speech disruption may occur, and higher frequency sounds), and metrics that align with critical human activities (e.g., Lnight to inform sleep disruption). Furthermore, while current geospatial modeling focuses on terrestrial surface noise, similar future work may advance efforts to map sounds in aquatic and other ecosystems.

## 4. Conclusion

The comparison between the BTS 2018 and 2020 national noise models indicated that noise levels between years were highly correlated, with an average nationwide difference of less than 1 dBA Leq, though greater differences exist in some regions of the US. Greater differences were observed between the BTS and NPS sound pressure level model results, even after conversion of the NPS model’s L50 sound level metric to Leq. Notably, transportation-related noise may be underestimated by the NPS model compared to the BTS model, but the NPS model may be useful in quantifying sound levels farther from transportation sources, which are censored in the BTS modeling results. We demonstrated that a hybrid contiguous national noise model that combines both BTS and NPS models has useful characteristics that address the inherent limitations of each individual model. When evaluated against sound level measurement data, the hybrid model showed good performance, with <1 dBA average bias and RMSE values of 4.9 dBA for metropolitan sites and 6.5 dBA for national park sites. The hybrid model is a useful representation of longer-term average sound levels, making it applicable for a variety of noise-related planning, policy, and research applications.

## Data Availability

All data produced in the present study are available upon reasonable request to the authors.

## Acknowledgements

We thank the NPS for providing us access to sound level monitoring data. We thank SFO, LAX, ORD and The Port Authority of New York and New Jersey for making noise data available on their websites.

