## Supplemental Information for "Comparison of Two National Noise Models: Progress Towards an Integrated Noise Model for Environmental Health Research in the United States"

**Table S1.** Differences between Leq and Ldn under the assumption of equivalent noise throughout the 24 hours for 40, 60, and 80 dBA scenarios. The difference between Ldn and Leq under these assumptions is 6.4 dBA.

| Hour | Leq | Penalty | Hour | Leq | Penalty | Hour | Leq | Penalty |
|---|---|---|---|---|---|---|---|---|
| 0 | 40 | 10 | 0 | 60 | 10 | 0 | 80 | 10 |
| 1 | 40 | 10 | 1 | 60 | 10 | 1 | 80 | 10 |
| 2 | 40 | 10 | 2 | 60 | 10 | 2 | 80 | 10 |
| 3 | 40 | 10 | 3 | 60 | 10 | 3 | 80 | 10 |
| 4 | 40 | 10 | 4 | 60 | 10 | 4 | 80 | 10 |
| 5 | 40 | 10 | 5 | 60 | 10 | 5 | 80 | 10 |
| 6 | 40 | 10 | 6 | 60 | 10 | 6 | 80 | 10 |
| 7 | 40 | | 7 | 60 | | 7 | 80 | |
| 8 | 40 | | 8 | 60 | | 8 | 80 | |
| 9 | 40 | | 9 | 60 | | 9 | 80 | |
| 10 | 40 | | 10 | 60 | | 10 | 80 | |
| 11 | 40 | | 11 | 60 | | 11 | 80 | |
| 12 | 40 | | 12 | 60 | | 12 | 80 | |
| 13 | 40 | | 13 | 60 | | 13 | 80 | |
| 14 | 40 | | 14 | 60 | | 14 | 80 | |
| 15 | 40 | | 15 | 60 | | 15 | 80 | |
| 16 | 40 | | 16 | 60 | | 16 | 80 | |
| 17 | 40 | | 17 | 60 | | 17 | 80 | |
| 18 | 40 | | 18 | 60 | | 18 | 80 | |
| 19 | 40 | | 19 | 60 | | 19 | 80 | |
| 20 | 40 | | 20 | 60 | | 20 | 80 | |
| 21 | 40 | | 21 | 60 | | 21 | 80 | |
| 22 | 40 | 10 | 22 | 60 | 10 | 22 | 80 | 10 |
| 23 | 40 | 10 | 23 | 60 | 10 | 23 | 80 | 10 |
| 24LAeq | 40 | | 24LAeq | 60 | | 24LAeq | 80 | |
| Ldn | | 46.4 | Ldn | | 66.4 | Ldn | | 86.4 |
| Difference | | 6.4 | | | 6.4 | | | 6.4 |

**Table S2.** Linear regression model of the relationships between 24-hour equivalent LEQ (dBA) (the regression outcome) and summer daytime L50 (dBA) (Model A).

| Characteristic | Beta | 95% CI | p-value |
|---|---|---|---|
| (Intercept) | 13.8 | 11.9, 15.8 | <0.001 |
| Summer daytime L50 | 0.770 | 0.717, 0.824 | <0.001 |

**Table S3.** Linear regression model of the relationship between 24-hour LEQ (dBA) (the regression outcome) and L50 (dBA) for all available seasons (Model B)

| Characteristic | Beta | 95% CI | p-value |
|---|---|---|---|
| (Intercept) | 13.6 | 12.2, 14.9 | <0.001 |
| L50 | 0.790 | 0.751, 0.829 | <0.001 |

**Table S4.** Pearson's and Spearman's correlation coefficients and average bias between BTS National Transportation Noise Model years 2018 and 2020, and the NPS National Sound Pressure Level Model by CBSA.

```
---------------------------------------
New York-Newark-Jersey City, NY-NJ
---------------------------------------
Pearson r:
         BTS_2018 BTS_2020  NPS
BTS_2018     1.00     0.93 0.13
BTS_2020     0.93     1.00 0.07
NPS          0.13     0.07 1.00

Spearman rho:
         BTS_2018 BTS_2020  NPS
BTS_2018     1.00     0.87 0.17
BTS_2020     0.87     1.00 0.11
NPS          0.17     0.11 1.00

BTS2020 - BTS2018: -1.01 (dBA)
BTS2020 - NPS: 2.22 (dBA)
BTS2018 - NPS: 2.13 (dBA)
---------------------------------------

---------------------------------------
Los Angeles-Long Beach-Anaheim, CA
---------------------------------------
Pearson r:
         BTS_2018 BTS_2020  NPS
BTS_2018     1.00     0.88 0.04
BTS_2020     0.88     1.00 0.00
NPS          0.04     0.00 1.00

Spearman rho:
         BTS_2018 BTS_2020  NPS
BTS_2018     1.00     0.85 0.09
BTS_2020     0.85     1.00 0.05
NPS          0.09     0.05 1.00

BTS2020 - BTS2018: 1.58 (dBA)
BTS2020 - NPS: 3.06 (dBA)
BTS2018 - NPS: 1.38 (dBA)
---------------------------------------

---------------------------------------
Chicago-Naperville-Elgin, IL-IN
---------------------------------------
Pearson r:
         BTS_2018 BTS_2020  NPS
BTS_2018     1.00     0.90 0.03
```

```
BTS_2020     0.90     1.00 0.11
NPS          0.03     0.11 1.00

Spearman rho:
         BTS_2018 BTS_2020  NPS
BTS_2018     1.00     0.87 0.07
BTS_2020     0.87     1.00 0.15
NPS          0.07     0.15 1.00

BTS2020 - BTS2018: 0.44 (dBA)
BTS2020 - NPS: 1.46 (dBA)
BTS2018 - NPS: 1.14 (dBA)
---------------------------------------

---------------------------------------
Dallas-Fort Worth-Arlington, TX
---------------------------------------
Pearson r:
         BTS_2018 BTS_2020  NPS
BTS_2018     1.00     0.93 0.09
BTS_2020     0.93     1.00 0.14
NPS          0.09     0.14 1.00

Spearman rho:
         BTS_2018 BTS_2020  NPS
BTS_2018     1.00     0.90 0.13
BTS_2020     0.90     1.00 0.19
NPS          0.13     0.19 1.00

BTS2020 - BTS2018: 0.31 (dBA)
BTS2020 - NPS: 1.58 (dBA)
BTS2018 - NPS: 1.55 (dBA)
---------------------------------------

---------------------------------------
Houston-Pasadena-The Woodlands, TX
---------------------------------------
Pearson r:
         BTS_2018 BTS_2020  NPS
BTS_2018     1.00     0.92 0.09
BTS_2020     0.92     1.00 0.11
NPS          0.09     0.11 1.00

Spearman rho:
         BTS_2018 BTS_2020  NPS
BTS_2018     1.00     0.89 0.16
BTS_2020     0.89     1.00 0.17
NPS          0.16     0.17 1.00

BTS2020 - BTS2018: -0.25 (dBA)
```

```
BTS2020 - NPS: 1.28 (dBA)
BTS2018 - NPS: 1.36 (dBA)
---------------------------------------

---------------------------------------
Atlanta-Sandy Springs-Roswell, GA
---------------------------------------
Pearson r:
         BTS_2018 BTS_2020  NPS
BTS_2018     1.00     0.90 0.24
BTS_2020     0.90     1.00 0.12
NPS          0.24     0.12 1.00

Spearman rho:
         BTS_2018 BTS_2020  NPS
BTS_2018     1.00     0.87 0.24
BTS_2020     0.87     1.00 0.13
NPS          0.24     0.13 1.00

BTS2020 - BTS2018: -1.12 (dBA)
BTS2020 - NPS: 2.64 (dBA)
BTS2018 - NPS: 3.05 (dBA)
---------------------------------------

---------------------------------------
Washington-Arlington-Alexandria, DC-VA-MD-WV
---------------------------------------
Pearson r:
         BTS_2018 BTS_2020  NPS
BTS_2018      1.0     0.90 0.10
BTS_2020      0.9     1.00 0.05
NPS           0.1     0.05 1.00

Spearman rho:
         BTS_2018 BTS_2020  NPS
BTS_2018     1.00     0.87 0.18
BTS_2020     0.87     1.00 0.08
NPS          0.18     0.08 1.00

BTS2020 - BTS2018: -0.54 (dBA)
BTS2020 - NPS: 3.35 (dBA)
BTS2018 - NPS: 3.02 (dBA)
---------------------------------------

---------------------------------------
Philadelphia-Camden-Wilmington, PA-NJ-DE-MD
---------------------------------------
Pearson r:
         BTS_2018 BTS_2020   NPS
BTS_2018     1.00     0.93 -0.01
```

```
BTS_2020     0.93     1.00 -0.02
NPS         -0.01    -0.02  1.00

Spearman rho:
         BTS_2018 BTS_2020   NPS
BTS_2018     1.00     0.92  0.00
BTS_2020     0.92     1.00 -0.01
NPS          0.00    -0.01  1.00

BTS2020 - BTS2018: -0.69 (dBA)
BTS2020 - NPS: 2.34 (dBA)
BTS2018 - NPS: 2.36 (dBA)
---------------------------------------

---------------------------------------
Miami-Fort Lauderdale-West Palm Beach, FL
---------------------------------------
Pearson r:
         BTS_2018 BTS_2020  NPS
BTS_2018     1.00     0.96 0.12
BTS_2020     0.96     1.00 0.06
NPS          0.12     0.06 1.00

Spearman rho:
         BTS_2018 BTS_2020  NPS
BTS_2018     1.00     0.94 0.21
BTS_2020     0.94     1.00 0.14
NPS          0.21     0.14 1.00

BTS2020 - BTS2018: -0.43 (dBA)
BTS2020 - NPS: -0.22 (dBA)
BTS2018 - NPS: 0.03 (dBA)
---------------------------------------

---------------------------------------
Phoenix-Mesa-Chandler, AZ
---------------------------------------
Pearson r:
         BTS_2018 BTS_2020   NPS
BTS_2018     1.00     0.93 -0.09
BTS_2020     0.93     1.00  0.06
NPS         -0.09     0.06  1.00

Spearman rho:
         BTS_2018 BTS_2020   NPS
BTS_2018     1.00     0.92 -0.02
BTS_2020     0.92     1.00  0.10
NPS         -0.02     0.10  1.00

BTS2020 - BTS2018: 0.31 (dBA)
```

```
BTS2020 - NPS: 5.01 (dBA)
BTS2018 - NPS: 3.52 (dBA)
---------------------------------------

---------------------------------------
Boston-Cambridge-Newton, MA-NH
---------------------------------------
Pearson r:
         BTS_2018 BTS_2020  NPS
BTS_2018     1.00     0.94 0.08
BTS_2020     0.94     1.00 0.04
NPS          0.08     0.04 1.00

Spearman rho:
         BTS_2018 BTS_2020  NPS
BTS_2018     1.00     0.94 0.08
BTS_2020     0.94     1.00 0.05
NPS          0.08     0.05 1.00

BTS2020 - BTS2018: -0.37 (dBA)
BTS2020 - NPS: 4.67 (dBA)
BTS2018 - NPS: 4.1 (dBA)
---------------------------------------

---------------------------------------
Riverside-San Bernardino-Ontario, CA
---------------------------------------
Pearson r:
         BTS_2018 BTS_2020   NPS
BTS_2018     1.00     0.89 -0.15
BTS_2020     0.89     1.00  0.05
NPS         -0.15     0.05  1.00

Spearman rho:
         BTS_2018 BTS_2020   NPS
BTS_2018     1.00     0.86 -0.14
BTS_2020     0.86     1.00  0.06
NPS         -0.14     0.06  1.00

BTS2020 - BTS2018: 2.63 (dBA)
BTS2020 - NPS: 8.49 (dBA)
BTS2018 - NPS: 6.26 (dBA)
---------------------------------------

---------------------------------------
San Francisco-Oakland-Fremont, CA
---------------------------------------
Pearson r:
         BTS_2018 BTS_2020  NPS
BTS_2018     1.00     0.86 0.08
```

```
BTS_2020     0.86     1.00 0.06
NPS          0.08     0.06 1.00

Spearman rho:
         BTS_2018 BTS_2020  NPS
BTS_2018     1.00     0.82 0.18
BTS_2020     0.82     1.00 0.14
NPS          0.18     0.14 1.00

BTS2020 - BTS2018: 1.06 (dBA)
BTS2020 - NPS: 2.69 (dBA)
BTS2018 - NPS: 1.54 (dBA)
---------------------------------------

---------------------------------------
Detroit-Warren-Dearborn, MI
---------------------------------------
Pearson r:
         BTS_2018 BTS_2020  NPS
BTS_2018     1.00     0.95 0.20
BTS_2020     0.95     1.00 0.17
NPS          0.20     0.17 1.00

Spearman rho:
         BTS_2018 BTS_2020  NPS
BTS_2018     1.00     0.92 0.22
BTS_2020     0.92     1.00 0.19
NPS          0.22     0.19 1.00

BTS2020 - BTS2018: -0.36 (dBA)
BTS2020 - NPS: 3.49 (dBA)
BTS2018 - NPS: 3.72 (dBA)
---------------------------------------

---------------------------------------
Seattle-Tacoma-Bellevue, WA
---------------------------------------
Pearson r:
         BTS_2018 BTS_2020  NPS
BTS_2018     1.00     0.96 0.02
BTS_2020     0.96     1.00 0.04
NPS          0.02     0.04 1.00

Spearman rho:
         BTS_2018 BTS_2020  NPS
BTS_2018     1.00     0.94 0.17
BTS_2020     0.94     1.00 0.13
NPS          0.17     0.13 1.00

BTS2020 - BTS2018: -0.53 (dBA)
```

```
BTS2020 - NPS: 1.63 (dBA)
BTS2018 - NPS: 1.54 (dBA)
---------------------------------------

---------------------------------------
Minneapolis-St. Paul-Bloomington, MN-WI
---------------------------------------
Pearson r:
         BTS_2018 BTS_2020  NPS
BTS_2018     1.00     0.89 0.05
BTS_2020     0.89     1.00 0.03
NPS          0.05     0.03 1.00

Spearman rho:
         BTS_2018 BTS_2020  NPS
BTS_2018     1.00     0.85 0.08
BTS_2020     0.85     1.00 0.01
NPS          0.08     0.01 1.00

BTS2020 - BTS2018: 0.06 (dBA)
BTS2020 - NPS: 3.13 (dBA)
BTS2018 - NPS: 2.2 (dBA)
---------------------------------------

---------------------------------------
Tampa-St. Petersburg-Clearwater, FL
---------------------------------------
Pearson r:
         BTS_2018 BTS_2020  NPS
BTS_2018     1.00     0.97 0.08
BTS_2020     0.97     1.00 0.05
NPS          0.08     0.05 1.00

Spearman rho:
         BTS_2018 BTS_2020  NPS
BTS_2018     1.00     0.96 0.12
BTS_2020     0.96     1.00 0.09
NPS          0.12     0.09 1.00

BTS2020 - BTS2018: -0.23 (dBA)
BTS2020 - NPS: 2.32 (dBA)
BTS2018 - NPS: 2 (dBA)
---------------------------------------

---------------------------------------
San Diego-Chula Vista-Carlsbad, CA
---------------------------------------
Pearson r:
         BTS_2018 BTS_2020  NPS
BTS_2018     1.00     0.90 0.03
```

```
BTS_2020     0.90     1.00 0.11
NPS          0.03     0.11 1.00

Spearman rho:
         BTS_2018 BTS_2020  NPS
BTS_2018     1.00     0.87 0.18
BTS_2020     0.87     1.00 0.21
NPS          0.18     0.21 1.00

BTS2020 - BTS2018: 1.83 (dBA)
BTS2020 - NPS: 4.17 (dBA)
BTS2018 - NPS: 1.95 (dBA)
---------------------------------------

---------------------------------------
Denver-Aurora-Centennial, CO
---------------------------------------
Pearson r:
         BTS_2018 BTS_2020  NPS
BTS_2018     1.00     0.94 0.21
BTS_2020     0.94     1.00 0.15
NPS          0.21     0.15 1.00

Spearman rho:
         BTS_2018 BTS_2020  NPS
BTS_2018     1.00     0.92 0.23
BTS_2020     0.92     1.00 0.17
NPS          0.23     0.17 1.00

BTS2020 - BTS2018: -0.32 (dBA)
BTS2020 - NPS: 4.75 (dBA)
BTS2018 - NPS: 4.69 (dBA)
---------------------------------------

---------------------------------------
Baltimore-Columbia-Towson, MD
---------------------------------------
Pearson r:
         BTS_2018 BTS_2020   NPS
BTS_2018     1.00     0.94  0.07
BTS_2020     0.94     1.00 -0.01
NPS          0.07    -0.01  1.00

Spearman rho:
         BTS_2018 BTS_2020  NPS
BTS_2018     1.00     0.92 0.15
BTS_2020     0.92     1.00 0.04
NPS          0.15     0.04 1.00

BTS2020 - BTS2018: -0.58 (dBA)
```

BTS2020 - NPS: 2.68 (dBA)
BTS2018 - NPS: 2.7 (dBA)
---------------------------------------

**Table S5. Estimated percentage of individuals within each age category across sound level exposure bins (dBA Leq).**

| dBA | Age Categories, % in age category | | | | | | | | |
|---|---|---|---|---|---|---|---|---|---|
| | All Ages | under 5 years | 5 to 17 years | 18 to 24 years | 25 to 44 years | 45 to 64 years | 65 to 74 years | 75 to 84 years | 85+ years |
| <35 | 0.10% | 0.09% | 0.10% | 0.09% | 0.09% | 0.10% | 0.11% | 0.11% | 0.07% |
| 35-39 | 1.26% | 1.09% | 1.22% | 0.93% | 1.06% | 1.35% | 1.84% | 1.66% | 1.19% |
| 40-44 | 12.72% | 11.67% | 12.72% | 10.30% | 10.77% | 14.12% | 16.13% | 15.48% | 12.36% |
| 45-49 | 28.69% | 28.33% | 30.19% | 26.99% | 26.47% | 29.67% | 30.37% | 30.67% | 29.77% |
| 50-54 | 46.17% | 47.64% | 46.01% | 48.38% | 48.25% | 44.78% | 42.63% | 43.28% | 46.94% |
| 55-59 | 9.51% | 9.61% | 8.32% | 11.50% | 11.58% | 8.57% | 7.61% | 7.49% | 8.26% |
| 60-64 | 0.90% | 0.91% | 0.85% | 1.09% | 1.01% | 0.82% | 0.76% | 0.74% | 0.80% |
| 65-69 | 0.36% | 0.37% | 0.32% | 0.40% | 0.42% | 0.33% | 0.30% | 0.30% | 0.33% |
| 70-74 | 0.15% | 0.15% | 0.15% | 0.16% | 0.17% | 0.15% | 0.13% | 0.15% | 0.14% |
| 75-79 | 0.09% | 0.08% | 0.08% | 0.09% | 0.10% | 0.08% | 0.08% | 0.08% | 0.10% |
| 80-84 | 0.04% | 0.04% | 0.04% | 0.05% | 0.05% | 0.03% | 0.03% | 0.02% | 0.03% |
| 85-89 | 0.01% | 0.01% | 0.00% | 0.01% | 0.01% | 0.01% | 0.00% | 0.01% | 0.00% |
| 90+ | 0.00% | 0.01% | 0.00% | 0.01% | 0.00% | 0.01% | 0.00% | 0.00% | 0.00% |

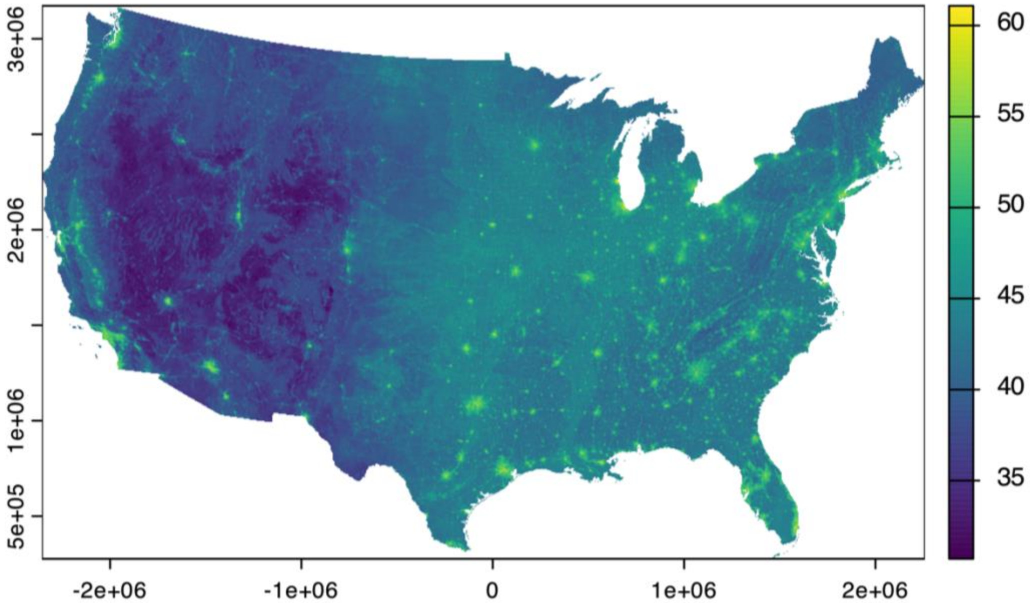

**Figure S1.** NPS national sound pressure level model converted to Leq (dBA) sound levels using Model A

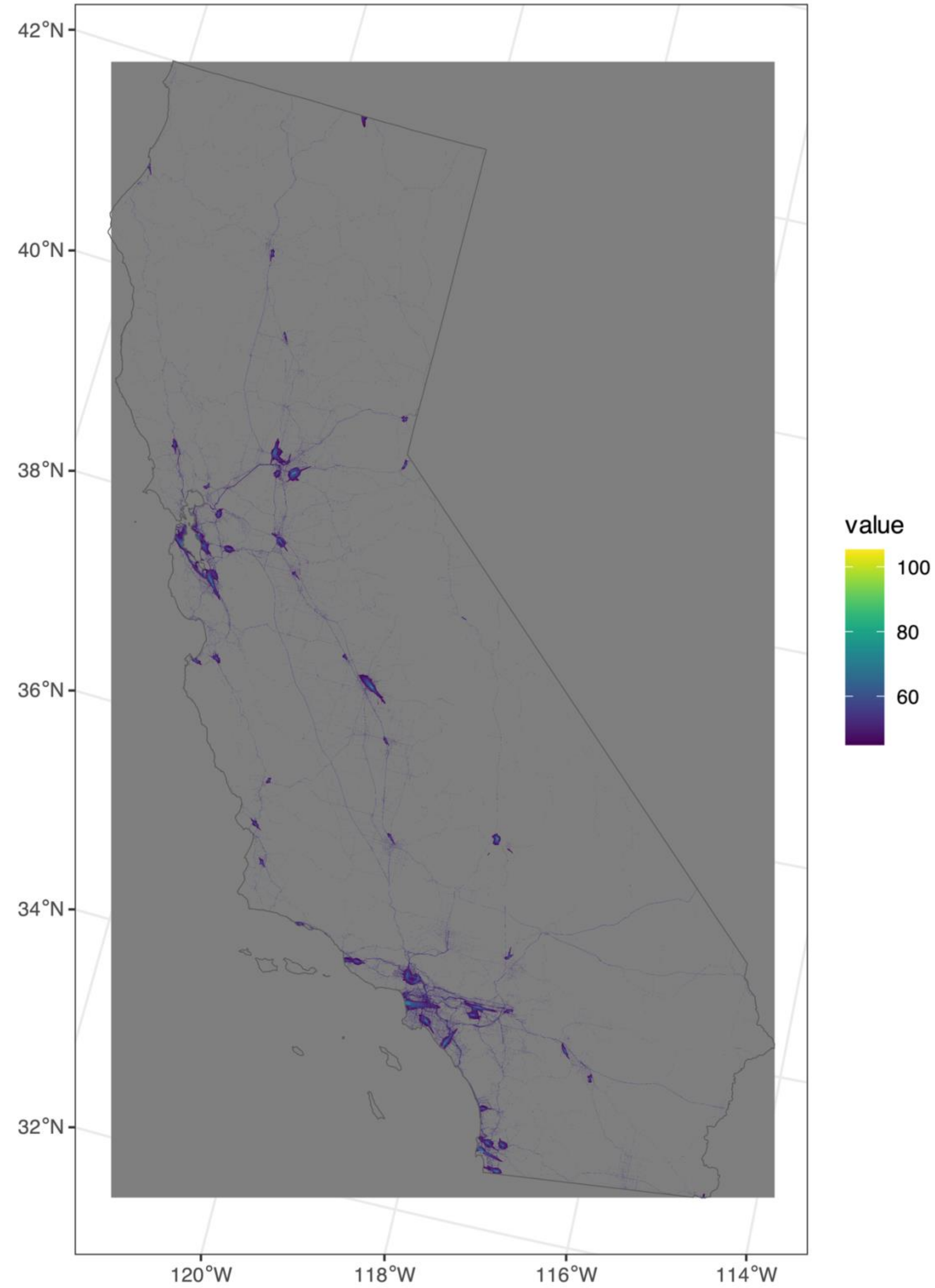


**Figure S2.** BTS national transportation noise map for the year 2018 showing Leq (dBA). Outline of the California state boundary is shown. Gray areas indicate areas with transportation noise levels below the 45 dBA model results threshold.

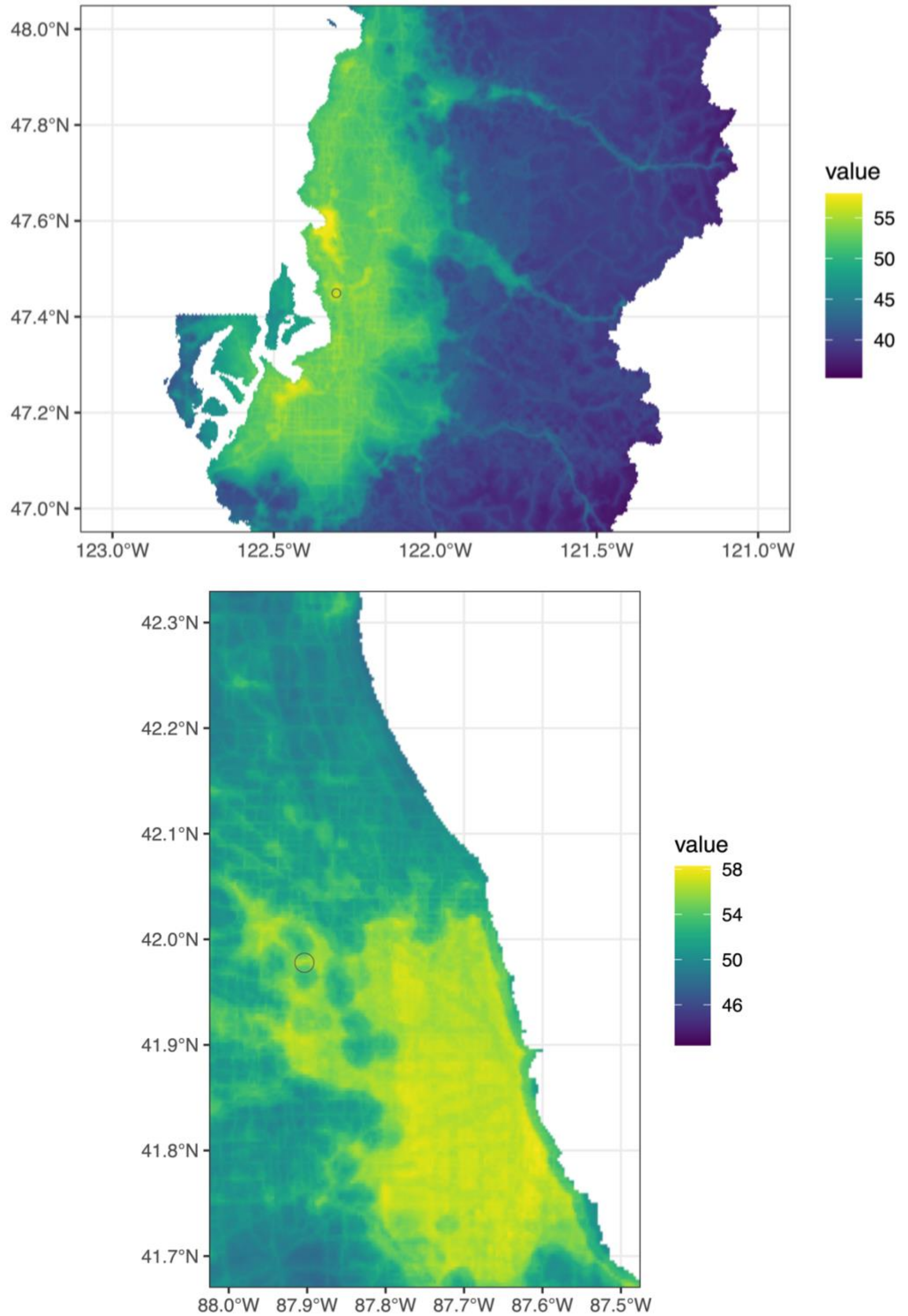


**Figure S3**. NPS sound pressure level model with converted Leq (dBA) values for an area of the Seattle, King County, WA CBSA, with a 1 km buffer shown for the Seattle-Tacoma International airport (top) and for the Chicago, IL CBSA, with a 1 km buffer shown for the Chicago O'Hare International Airport (bottom).

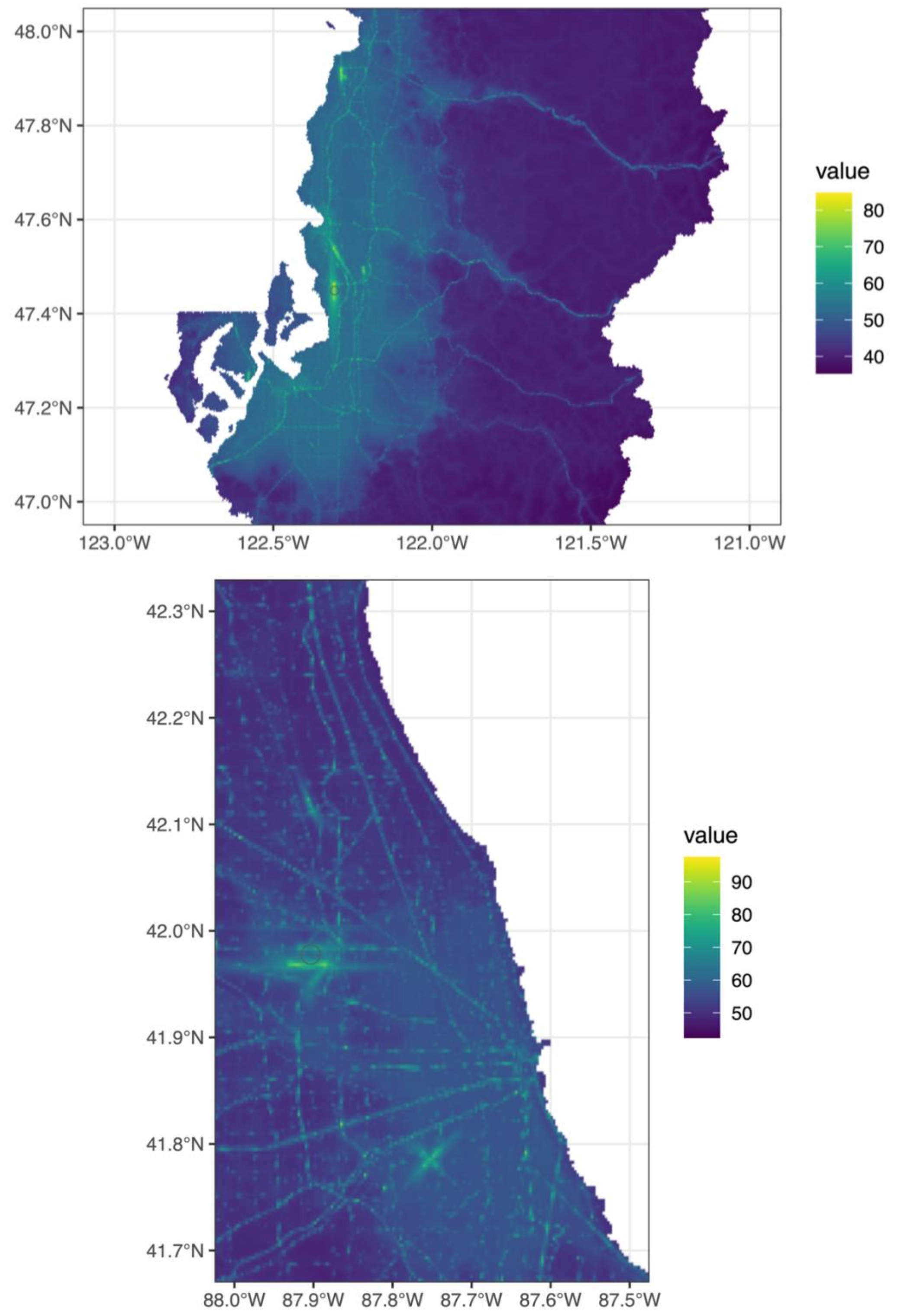


**Figure S4.** Hybrid model Leq (dBA) values for an area of the Seattle, King County, WA CBSA, with a 1 km buffer shown for the Seattle-Tacoma International Airport (top) and for the Chicago, IL CBSA, with a 1 km buffer shown for the Chicago O'Hare International Airport (bottom).
